# Abnormal lymphatic S1P signaling aggravates lymphatic dysfunction and tissue inflammation

**DOI:** 10.1101/2023.06.08.23291175

**Authors:** Dongeon Kim, Wen Tian, Timothy Ting-Hsuan Wu, Menglan Xiang, Ryan Vinh, Jason Chang, Shenbiao Gu, Seunghee Lee, Yu Zhu, Torrey Guan, Emilie Claire Schneider, Evan Bao, J. Brandon Dixon, Peter Kao, Junliang Pan, Stanley G. Rockson, Xinguo Jiang, Mark Robert Nicolls

**Affiliations:** VA Palo Alto Health Care System, Palo Alto, California, USA; Stanford University School of Medicine, Stanford, California, USA; Department of Biochemistry, Stanford Bio-X, Stanford, California, USA; Georgia Institute of Technology, Atlanta, Georgia, USA

**Keywords:** Lymphedema, Lymphatic endothelial cells (LECs), Sphingosine-1-phosphate receptor 1 (S1PR1), P-selectin, and CD4 T cells

## Abstract

**BACKGROUND:** Lymphedema is a global health problem with no effective drug treatment. Enhanced T cell immunity and abnormal lymphatic endothelial cell (LEC) signaling are promising therapeutic targets for this condition. Sphingosine-1-phosphate (S1P) mediates a key signaling pathway required for normal LEC function, and altered S1P signaling in LECs could lead to lymphatic disease and pathogenic T cell activation. Characterizing this biology is relevant for developing much-needed therapies.

**METHODS:** Human and mouse lymphedema was studied. Lymphedema was induced in mice by surgically ligating the tail lymphatics. Lymphedematous dermal tissue was assessed for S1P signaling. To verify the role of altered S1P signaling effects in lymphatic cells, LEC-specific *S1pr1*-deficient (*S1pr1*^LECKO^) mice were generated. Disease progression was quantified by tail-volumetric and -histopathological measurements over time. LECs from mice and humans, with S1P signaling inhibition, were then co-cultured with CD4 T cells, followed by an analysis of CD4 T cell activation and pathway signaling. Finally, animals were treated with a monoclonal antibody specific to P-selectin to assess its efficacy in reducing lymphedema and T cell activation.

**RESULTS:** Human and experimental lymphedema tissues exhibited decreased LEC S1P signaling through S1PR1. LEC *S1pr1* loss-of-function exacerbated lymphatic vascular insufficiency, tail swelling, and increased CD4 T cell infiltration in mouse lymphedema. LECs, isolated from *S1pr1*^LECKO^ mice and co-cultured with CD4 T cells, resulted in augmented lymphocyte differentiation. Inhibiting S1PR1 signaling in human dermal LECs (HDLECs) promoted T helper type 1 and 2 (Th1 and Th2) cell differentiation through direct cell contact with lymphocytes. HDLECs with dampened S1P signaling exhibited enhanced P-selectin, an important cell adhesion molecule expressed on activated vascular cells. *In vitro*, P-selectin blockade reduced the activation and differentiation of Th cells co-cultured with sh*S1PR1*-treated HDLECs. P-selectin-directed antibody treatment improved tail swelling and reduced Th1/Th2 immune responses in mouse lymphedema

**CONCLUSION:** This study suggests that reduction of the LEC S1P signaling aggravates lymphedema by enhancing LEC adhesion and amplifying pathogenic CD4 T cell responses. P-selectin inhibitors are suggested as a possible treatment for this pervasive condition.

**Clinical Perspective:** *What is New?:*

- Lymphatic-specific *S1pr1* deletion exacerbates lymphatic vessel malfunction and Th1/Th2 immune responses during lymphedema pathogenesis.
- *S1pr1*-deficient LECs directly induce Th1/Th2 cell differentiation and decrease anti-inflammatory Treg populations.
- Peripheral dermal LECs affect CD4 T cell immune responses through direct cell contact.
- LEC P-selectin, regulated by S1PR1 signaling, affects CD4 T cell activation and differentiation.
- P-selectin blockade improves lymphedema tail swelling and decreases Th1/Th2 population in the diseased skin.

*What Are the Clinical Implications?:*

- S1P/S1PR1 signaling in LECs regulates inflammation in lymphedema tissue.
- S1PR1 expression levels on LECs may be a useful biomarker for assessing predisposition to lymphatic disease, such as at-risk women undergoing mastectomy
- P-selectin Inhibitors may be effective for certain forms of lymphedema

## Introduction

Lymphedema is a chronic state of lymphatic vascular insufficiency characterized by regionally impaired immunity, interstitial edema, adipose deposition, and fibrotic remodeling. The condition affects around 100-200 million individuals globally but has no effective pharmacological therapies ^1, 2^. Secondary lymphedema is the predominant form of the disease, resulting from acquired lymphatic vascular damage arising after parasitic infection, oncologic conditions, cancer therapy, chronic venous disease, and trauma. Research from the past two decades suggests that pathologically skewed CD4^+^ T cell differentiation promotes preclinical lymphedema ^3, 4^. While blocking Th2 immunity or inflammation, in general, improves skin histopathology, this therapy is ineffective for reducing limb volumes ^5^. These studies demonstrate the relevance of abnormal immunity to lymphedema in a way that may inform therapeutic design.

By producing molecules such as S1P, adhesion proteins, and CCL21, lymphatic endothelial cells (LECs) facilitate the lymphatic transport of tissue fluid, antigens, and antigen-presenting cells (APCs) to draining lymph nodes (LNs) ^6–8^. Emerging studies show that LECs are critical immune cell modulators ^9–11^. For example, LECs regulate T cell activation and immune tolerance in peripheral tissue through the programmed cell death protein-ligand inhibitory pathway and MHC I/II complexes ^9, 12–14^; LECs also inhibit the maturation of dendritic cells by dampening their ability to activate effector T cells ^15^. Unlike the better-understood roles of LEC-centered lymphangiogenesis and lymph trafficking in lymphedema, the action of these vascular cells in disease-associated inflammation is less well understood.

Sphingosine-1-phosphate (S1P) is a bioactive sphingolipid metabolite involved in many cellular processes, including differentiation, survival, migration, angiogenesis, and lymphangiogenesis ^16–19^. It exerts biological effects through binding and activating five closely related G-protein coupled receptors, namely, S1P receptor (S1PR) 1, 2, 3, 4, and 5 ^20^. S1P signaling, via S1PR1 in vascular endothelial cells, promotes restoration of vascular integrity and induces cell-cell adhesion between pericytes and endothelial cells ^21–24^. S1PR1 is the most highly expressed isoform in LECs and is a necessary receptor to prevent lymphatic hypersprouting and promote lymphatic maturation ^18, 25, 26^. While various types of cells synthesize S1P, endothelial cells, including blood and lymphatic endothelial cells, are generally considered as major producers ^18^. S1P generation is controlled by sphingosine kinases SPHK1 and SPHK2, with the former enzyme playing the more dominant role. A prior study shows that SPHK1 expression and the lyase that degrades S1P are significantly downregulated in mouse lymphedema ^27^, raising the possibility that S1P signaling abnormality may promote lymphedema development.

In this study, we showed that LEC S1PR1 expression was low in both clinical and pre-clinical lymphedema skin. Using LEC-specific *S1pr1* loss-of-function transgenic mouse lines (*S1pr1*^LECKO^), we found that lymphatic *S1pr1* deficiency exacerbates lymphedema. Cell culture studies demonstrated that LECs with reduced S1PR1 skewed T cell differentiation towards T helper 1/2 (Th1/Th2) phenotypes in a contact-dependent manner. Bulk mRNA-Sequencing analysis revealed that S1P signaling deficiency enhanced LEC expression of P-selectin, an adhesion molecule regulating T cell trafficking and activity ^28^. Blocking P-selectin attenuated Th1/Th2 differentiation induced by *S1PR1*-deficient LECs in culture; anti-P-selectin antibody treatment decreased tail swelling and reduced Th1/Th2 cell population in lymphedema skin of *S1pr1*^LECKO^ mice. Collectively, we describe how lymphedema-associated immune dysregulation may be linked to reduced LEC S1P signaling and how P-selectin inhibitors, already approved for use in other diseases, may be effective in this otherwise refractory condition.

## Methods

All data and methods used in analysis will be made available to any researcher upon reasonable request.

### Mice

All mice were purchased from Jackson Laboratory. Detailed mice information: C57BL/6J (B6, JAX:000664), Prox1tm3(cre/ERT2)Gco/J (Prox1-Cre-ER^T2^, JAX:022075), B6.129S6(FVB)-S1pr1tm2.1Rlp/J (S1P ^loxp^, JAX:019141), and B6.Cg *Gt(ROSA)26Sor^tm^*^14(CAG–tdTomato)^*^Hze^*/J (Ai14(RCL-tdT)-D, JAX:007914) To generate lymphatic endothelial-specific *S1pr1*-deficient transgenic strain, mice expressing *Prox1-Cre-ER^T^*^2^ were crossed with *S1PR1^loxp^* mice. *Prox1-Cre-ER^T^*^2^ alone littermates were used as WT controls. To generate lymphatic endothelial-specific *tdTomato* reporter mice (*Prox1-Cre-ER^T^*^2^*-tdTomato*), *Prox1-Cre-ER^T^*^2^ mice were crossed with *tdTomato* mice. 250mg/kg tamoxifen was subcutaneously injected to 8 weeks mice for 3 consecutive days to activate the Cre-loxP system.

### Study approval

Investigation of human tissues in this study was approved by the Stanford University Institutional Review Board (IRB Protocol 7781). The samples were derived from adult patients with chronic primary and acquired lymphedema. Tissue specimens consisted of two contiguous 6-mm full-thickness cutaneous punch biopsy specimens obtained from a lymphedema-affected limb, with similar specimens obtained from the unaffected contralateral limb serving as control specimens. The diagnosis of lymphedema for each subject was clinically ascertained in the Stanford Center for Lymphatic and Venous Disorders. Phlebotomy samples for quantitation of serum S1P levels were obtained from lymphedema subjects and healthy controls; studies using these phlebotomy specimens were approved by the Stanford University Institutional Review Board (IRB Protocol 17690). Human buffy coats were purchased from Stanford Blood Center. All animal studies were approved by the VA Palo Alto Institutional Animal Care and Use Committee (IACUC).

### The mouse-tail model of acquired lymphedema

Lymphedema was induced through ablation of major lymphatic trunks on both sides of the tail and dermal lymphatic capillaries. Tail skin incision was made ∼2 cm from the mouse tail base. For surgical sham controls, skin incision alone was performed. Disease progression was quantified by volumetric and histopathological measurements. Lymphatic vessel functions were analyzed with a near-infrared (NIR) imaging system.

### Tail volume measurement

Mice tail images were taken through digital photographic technique preoperatively (D0) and postoperatively (D7, 14, and 21) using an Olympus D-520 Zoom digital camera at a fixed distance from the subject (37cm). Digital photographic tail images were quantified using Adobe Photoshop ruler tool. Tail volumes were derived from measurement of the tail diameter using the truncated cone approximation method **(Figure S1)**.

### Quantification of S1P with liquid chromatography tandem mass spectrometry (LC-MS/MS)

Briefly, S1P was separated from serum with an ACE reverse phase C18 HPLC column. Formic acid 0.5% in 5mM NH_4_Ac (A) and formic acid 0.5% in acetonitrile/water (9/1) (B) were used as the mobile phase. Lipids were detected with positive multiple reaction monitor (MRM) scanning at 330.20/264.40 m/z for S1P using a Sciex API-4000 MS/MS combined with a Shimazu 20A HPLC system. The internal standard for this method was Carbutamide. The quantification limit was 1 ng/ml. The calibration range was 1-2000ng/ml. The accuracy of the standards and control samples ranged from 88.7% to 120%. The information of patients was described in **Table S1 and S2**.

### Lymphatic drainage and leakiness test by NIR imaging

The lymphatic vessel transportation function and leakage were characterized by using a NIR lymphatic imaging system and dye quantification at day 21. The collecting lymphatic function was tracked throughout the procedure by imaging the dynamics of transport of a 10 µl ICG (10mM) injected intradermally, at the tip of the mouse tail. NIR images were taken 10 minutes after dye injection using a custom NIR imaging system as described in detail in the previous study ^29, 30^. ImageJ was used for the quantification of NIR dye leakage. Specifically, the relative intensity of leaked ICG was determined in two steps: 1) total leaked NIR intensity from each tail was calculated and averaged; 2) intensity was then normalized to control (which was set to 1).

### Embryo lymphatic development study

2 mg tamoxifen was injected intraperitoneally (i.p.) to pregnant mice (*Prox-1-Cre^ERT2^* x *S1pr1^fl/-^* mated with *Prox-1-Cre^ERT2^* x *S1pr1^fl/-^*) at E10.5 and E11.5 as previously described ^29^. Embryos were harvested at E17.5. Brightfield images were taken by using MVX10 microscope. Dorsal areas of embryonic skin were harvested for lymphatic vascular evaluation.

### Immunofluorescence (IF) staining

Surgery sites of 10 μm tail frozen sections were used for immunohistochemistry. Tail tissues were snap-frozen in O.C.T. compound solution (Fisher Healthcare) for IF staining and paraffin-embedded for hematoxylin and eosin (H&E) staining. Anti-LYVE1 (1:50; LSBio) Ab for lymphatic vessels and anti-CD4 (1:50; Abcam) Ab for CD4 T cell were stained at 4℃ overnight. Secondary antibodies were labeled with the Alexa Fluor 488 or Cy3 for 1h at room temperature (1:400; Invitrogen). Photomicrographs were acquired using LEICA DMi8 or Zeiss LSM710. ImageJ was used to quantify for lymphatic areas.

### Real-time reverse transcription quantitative PCR (RT-qPCR)

Total RNA was isolated from mouse tail skin using the RNeasy® Fibrous Tissue Mini Kit (QIAGEN) and from HDLECs using RNeasy® Mini Kit (QIAGEN) according to the manufacturer’s instructions. Total RNA was synthesized to complementary DNA using the High-Capacity cDNA Reverse Transcription Kits (Appliedbiosystems) following the manufacturer’s protocol. RT-PCR was performed using PowerSYBR Green PCR Master Mix (Applied Biosystems) according to the manufacturer’s instructions. The amplification condition was set at 40 cycles at 95℃ for 15s and 60℃ for 60s using AMI Prism® 7900HT Sequence detection System (Applied Biosystems). The sequences of the primers used for RT-qPCR are listed in **Table S3 and S4**.

### CD4^+^ T cell isolation

Naïve CD4^+^ T cells from the spleens of C57BL/6 WT mice were purified with Naïve CD4^+^ T cell Isolation Kit according to the manufacturer’s (Miltenyi Biotec) instructions. Peripheral blood mononuclear cells (PBMCs) were purified from buffy coats by gradient centrifugation with histopaque (density: 1.077 g/ml). Naïve CD4 T cells were purified from PBMCs using EasySep Human Naïve CD4^+^ T cell Isolation Kit II (STEMCELL Technologies). Memory CD4 T cells were purified from PBMCs using EasySep Human Memory CD4^+^ T cell Isolation Kit (STEMCELL Technologies).

### LEC purification and culture

Axillary, bronchial, and linguinal LNs were isolated from WT and *S1pr1*^LECKO^ mice. LNs were digested with the multi-tissue dissociation Kit I and gentle MACS Octo Dissociator with Heaters (Miltenyi Biotec). Digested lymph node stromal cells were cultured in endothelial cell growth medium II supplemented with 2% fetal calf serum, 100 U/mL of penicillin, 100 g/mL of streptomycin, and growth supplements including epidermal growth factor (5ng/ml), basic fibroblast growth factor (10ng/ml), insulin-like growth factor (20 ng/ml), vascular endothelial growth factor 165 (0.5 ng/ml), heparin (22.5 μg/ml), and hydrocortisone (0.2 μg/ml). After five days of culture, cells of CD45^-^, CD31^+^, and Gp38^+^ were selected as LEC population. Cells were sorted by fluorescence-activated cell sorting (FACS) Aria III. LECs used in experiments showed over 99% purity.

### Co-culture of CD4^+^ T cells with LECs

Purified LECs were cultured in a 24-well plate (4 x 10^4^/well). After overnight adhesion, 2 x 10^5^ mouse naïve CD4^+^ T cells and APCs along with anti-CD3 and CD28 monoclonal antibodies (BD Biosciences; 3 µg/ml) were added and cultured for 3 days in RPMI-1640 supplemented with 10% FBS and 1% penicillin-streptomycin at 37℃ in a humidified 5% CO2 atmosphere. Commercially-purchased HDLECs (PromoCell) were cultured in a 24-well plate (4 x 10^4^/well). After overnight adhesion, 2 x 10^5^ human naïve CD4^+^ T cells with ImmunoCult^TM^ Human CD3/CD28 T cell Activator (STEMCELL Technologies) were added and cultured for 3 days in RPMI-1640 supplemented with 10% FBS and 1% penicillin-streptomycin at 37°C in a humidified 5% CO_2_ atmosphere.

### Flow cytometry analysis

Single-cell suspension prepared from mice tail skin digested with multi tissue dissociation Kit I or from *ex vivo* cultures were analyzed by intracellular cytokine staining as previously described ^31, 32^. Prepared cells were restimulated with Cell Stimulation Cocktail (plus protein transport inhibitors) (ThermoFisher Scientific) for 5 h at 37℃ in a humidified 5% CO_2_ atmosphere. After stimulation, the cells were stained with fluorescein-conjugated antibodies against Fixable Viability Dye (Invitrogen), CD4, CD8, CD25, and CD44 (BD Bioscience). Following surface staining, Foxp3, IFN-ɣ, IL-4, and IL-17A were intracellularly stained using fixation/permeabilization buffer (eBioscience) according to the manufacturer’s instructions. Additionally, CD31, CD45, CD69, CD103, Gp38, and S1PR1 fluorescein-conjugated antibodies were used for surface staining. Isotype-matched antibodies were used as isotype controls. **Table S5** summarize the Ab information.

### Cytokine profiling

The supernatant co-cultured human CD4 T cell with HDLEC was collected at day 3. T cell related-cytokine assay was performed in EVE technologies.

### Analysis of bulk mRNA sequencing (bulk RNA-seq) data

Total RNA was isolated from HDLECs using the RNeasy® Plus Mini Kit (QIAGEN) according to the manufacturer’s instructions. mRNA library was formed by ploy A enrichment and reverse transcription of cDNA. Illumina PE150 technology is processed to sequence the sample (Novogene). Sequencing reads were aligned to the Human GRCh38 reference genome according to the ENCODE uniform processing pipeline, by STAR (v2.1.3) then quantified with RSEM (v1.4.1). Differential gene expression analysis was performed in R using EBSeq package, an empirical Bayes hierarchical model for inference in RNA-seq experiments, using false discovery rate (FDR) of 0.05 to retrieve list of differentially expressed genes (DEGs). Differentially expressed genes were ranked by posterior fold change and enriched for gene sets using The Molecular Signatures Database (MSigDB) and Gene Set Enrichment Analysis (GSEA) tools developed by the Broad Institute.

### Statistics

All data are presented as a mean ± SEM. Nonparametric Mann-Whitney test and Wilcoxon matched-pairs signed rank test were used for statistical analyses using GraphPad Prism v9.3.1. For comparisons between multiple experimental groups, Ordinary one-way ANOVA followed by Dunn’s multiple comparisons test for post hoc analyses were used after normal distribution test. p < 0.05 was considered statistically significant. Error bars correspond to the mean with SEM.

## Results

### Lymphedema is characterized by decreased LEC S1PR1-mediated S1P signaling

LEC S1P signaling through S1PR1 is critical for lymphatic homeostasis and repair ^26^. To determine the relevance of LEC S1P/S1PR1 signaling in lymphedema, we used a mouse tail lymphedema model, in which tail skin incision and lymphatic trunk ablation were performed for surgical induction of lymphedema (**Figure 1A**). Real-time reverse transcription quantitative PCR (RT-qPCR) analysis of *Sphk1*, the key gene involved in S1P production, demonstrated a reduction of the expression in lymphedematous mouse skin (**Figure 1B**), confirming our prior microarray study ^27^. LECs are the major cell type in the skin that produces S1P, a process reliant on SPHK1 activity ^26, 33–35^. LECs in lymphedema tissues express decreased SPHK1 (**Figure 1C, 1D, and S2A)**. S1PR1 is the most strongly expressed S1P receptor in LECs, and S1PR1 signaling is necessary for lymphatic vessel maturation ^26^. A diminished *S1pr1* mRNA level was detected in lymphedema tail skin (**Figure 1E**). More specifically, we analyzed S1PR1 expression in Gp38^+^CD31^+^ cells using flow cytometry and found significantly decreased S1PR1 expression in LECs in lymphedema skin (**Figure 1F and 1G**). Next, we assessed S1P serum concentration in preclinical and clinical lymphedema. Decreased S1P production was detected in the lymphedema condition compared with the healthy control in both mouse and human (**Figure 1H, 1I, and S2B)**. Consistent with mouse lymphedema tail skin, the expression of S1PR1 was decreased in Gp38^+^ lymphatic vessel of human lymphedema skin (**Figure 1J and 1K**). These data demonstrate that LEC S1P/S1PR1 signaling is reduced in both human and preclinical lymphedema.

**Figure 1.**
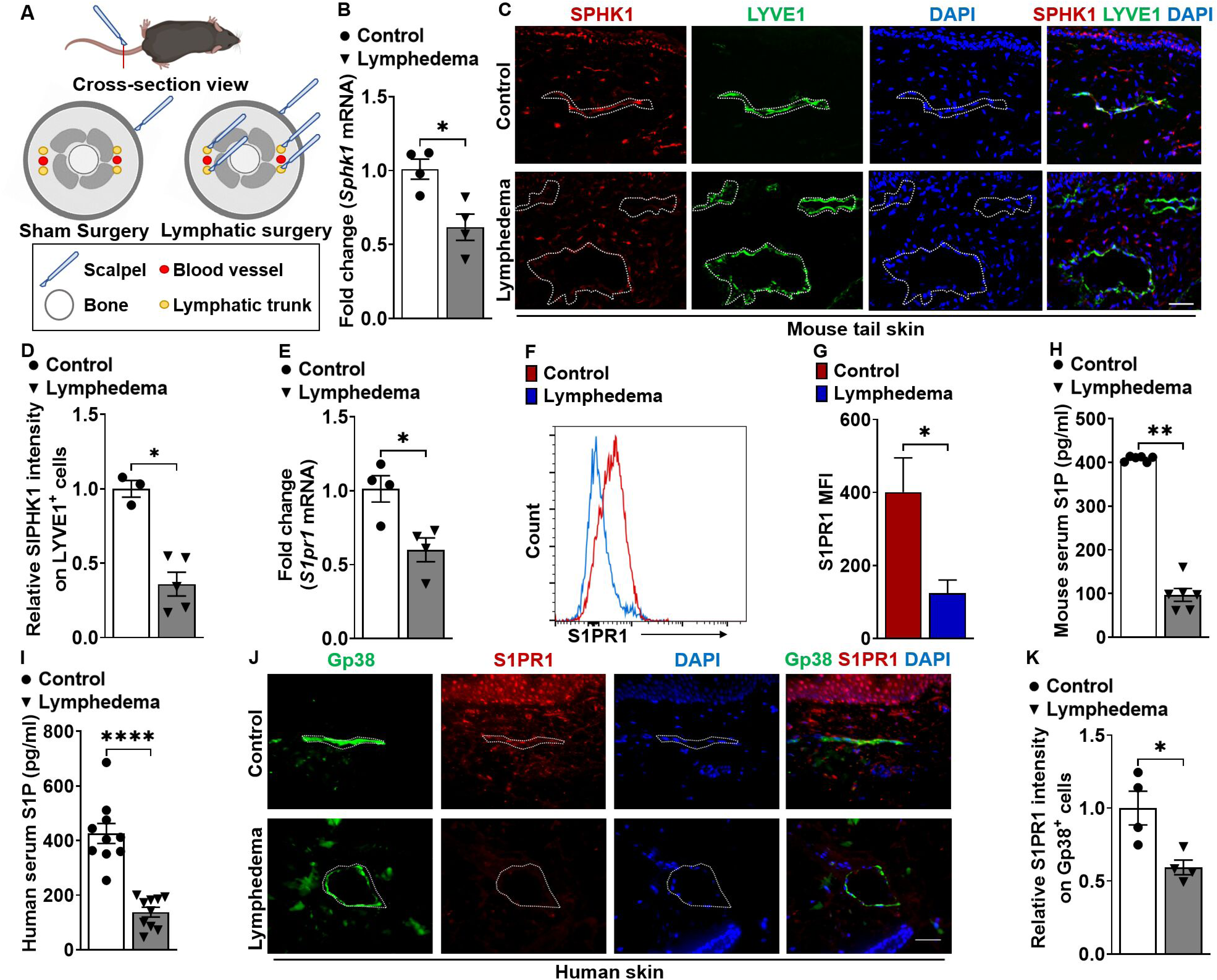
S1PR1 signaling of LECs is reduced in both mouse and human lymphedema skin. (**A**) Acquired lymphedema was surgically induced in the tails of C57BL/6J mice through thermal ablation of lymphatic trunks. Skin incision, alone, was performed in sham surgery groups. **(B)** RT-qPCR analysis of *Sphk1* mRNA levels in tail skin from control sham surgery mice or animals subjected to lymphatic surgery (n = 4 per each group). **(C)** Representative immunofluorescence (IF) staining of SPHK1 (red) and LYVE1 (green) of the skin tissues harvested from control or lymphedema mice. DAPI (blue) stains for the nucleus. Scale bar = 40 µm. **(D)** Quantification of the SPHK1 staining intensity comparing groups shown in C (n ≥ 3 per each group). **(E)** RT-qPCR analysis of *S1pr1* mRNA levels in tail skin from control sham surgery mice or lymphedema mice (n = 4 per each group). **(F and G)** Flow cytometry histograms show the mean fluorescence intensity of S1PR1 on LEC (Gp38^+^CD31^+^) population. Representative (F) and compiling data (G) are shown (n ≥ 3 per each group). **(H and I)** The serum concentration of S1P in mouse (H) and human (I) lymphedema (n = 10 per each group). **(J)** Representative IF staining of Gp38 (green) and S1PR1 (red) of the human skin from healthy control or lymphedema. DAPI (blue) stains for the nucleus. Scale bar = 50 µm. **(K)** Quantification of the S1PR1 intensity comparing groups shown in J (n = 4 per each group). Data are presented as mean ± SEM; * p < 0.05, ** p < 0.01, and **** p < 0.0001 by the Mann-Whitney test.

### Lymphatic *S1pr1* deficiency promotes lymphatic dysfunction in lymphedema

To evaluate lymphatic-specific S1P/S1PR1 signaling in lymphedema pathogenesis, we generated LEC-specific *S1pr1*-deficient (*S1pr1*^LECKO^) mice by crossing *S1pr1*^fl/fl^ mice with transgenic mice expressing Cre-ERT2 under the control of the Prox-1 promoter (*Prox-1-Cre^ERT2^*). Assessment of LEC (Gp38^+^CD31^+^) S1PR1 expression demonstrated >75% knockout efficiency **(Figure S3)**. *S1pr1*^LECKO^ mice were subjected to lymphedema surgery three weeks following tamoxifen treatment (**Figure 2A**). Serial evaluation of tail volumes following lymphedema surgery showed that genetic silencing of LEC-specific *S1pr1* exacerbated tail swelling compared with WT littermate control mice (**Figure 2B and 2C**). Sham surgery, which only involves skin incision, did not cause tail swelling in both WT and *S1pr1*^LECKO^ mice **(Figure S4)**. The mouse lymphedema model is characterized by severe cutaneous thickening and the lymphatic area increase in affected tails (**Figure 2D**). H&E staining of cross-sections demonstrated increased cutaneous thickness and lymphatic area in tissues derived from *S1pr1*^LECKO^ mice (**Figure 2E through 2G)**. Immunofluorescence (IF) staining similarly demonstrated areas of increased lymphatic capillaries identified by LYVE1 positivity in tail samples from *S1pr1*^LECKO^ mice (**Figure 2H and 2I**). Additionally, proliferation marker KI-67^+^ cells in LYVE-1^+^ cells were elevated in lymphatic *S1pr1*-deficient mice with lymphedema comparing to that of the WT control, suggesting that augmented number of LECs contributes to the increase of LYVE1^+^ lymphatic vascular area **(Figure S5)**.

**Figure 2.**
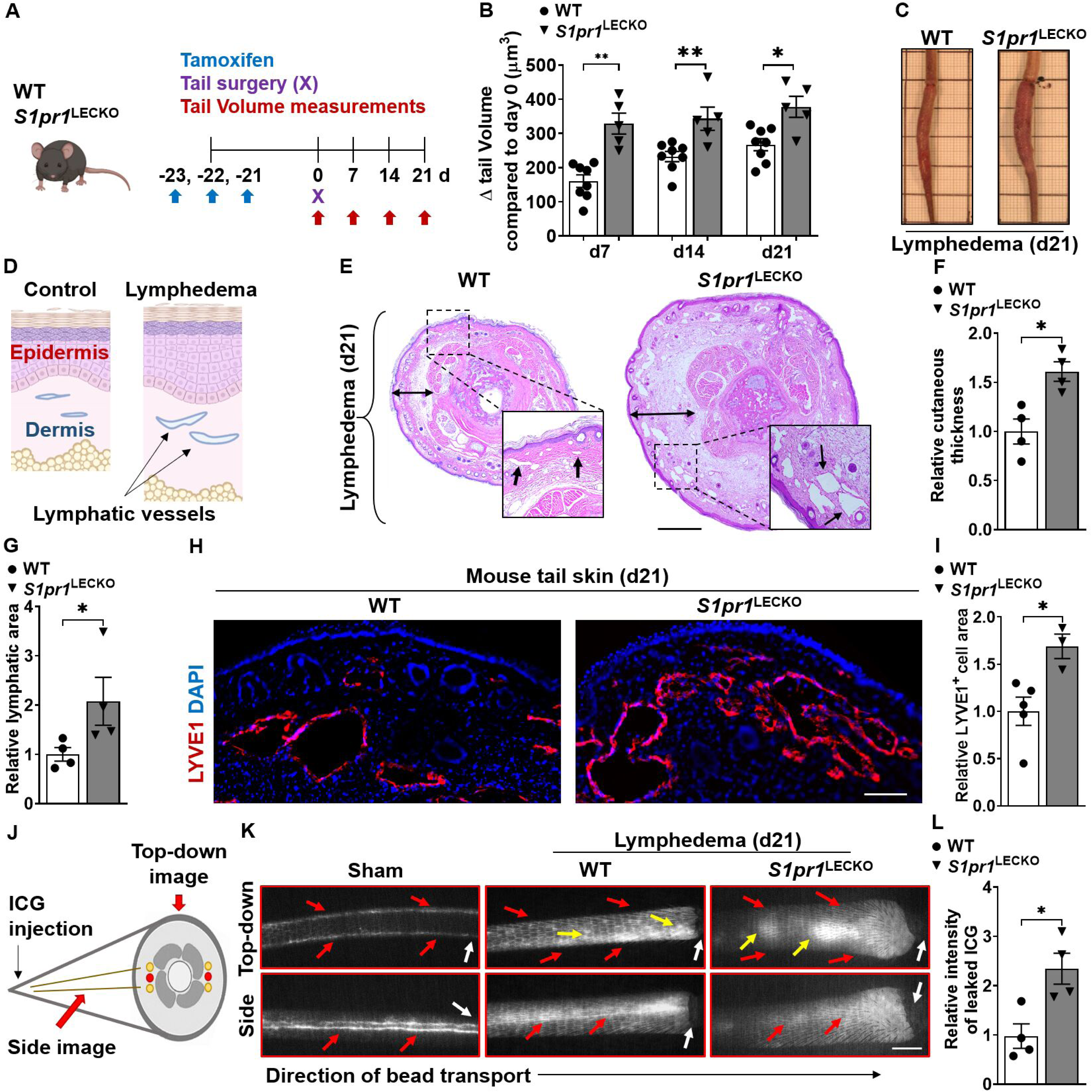
LEC-specific *S1pr1* deficiency exaggerates tissue swelling, augments cutaneous skin thickness, promotes lymphatic leakage, and reduces lymphatic drainage. **(A)** Schematic diagram of the experimental protocol. **(B)** Quantification of tail volume changes over time of WT and *S1pr1*^LECKO^ mice after lymphatic surgery (n ≥ 5 per each group). **(C)** Representative photographs of tails 21d following lymphatic surgery. **(D)** A cartoon showing the cross-section view of mouse-tail skin with or without lymphedema. **(E toG)** H&E staining of tail cross-section 21d following lymphatic surgery. Representative images (E) are shown. Black arrows indicate lymphatic vessel areas. Double-headed black arrows demonstrate cutaneous thickness. Quantification of cutaneous thickness (F) and lymphatic vessel luminal areas (G) are shown (n = 4 per each group). Scale bar = 1 mm. **(H and I)** Immunofluorescent images of LYVE1 (red) of tail skin 21d after surgery. Representative image (H) and quantification of the LYVE1 area data (I) are shown (n ≥ 3 per each group). DAPI (blue) stains the nucleus. Scale bar = 100 µm. **(J)** A cartoon showing one side of the mouse tail with two-line lymphatic trunks. **(K and L)** Near-infrared imaging 21d following lymphatic surgery after ICG injection near the tip of the tail. Representative images (K) are shown. White arrows denote surgical sites. Red arrows indicate ICG drainage. Yellow arrows indicate ICG leakage. Scale bar = 0.5 cm. Quantification of leakage (L) is shown (n = 4 per each group). Data are presented as the mean ± SEM; * p < 0.05 and ** p < 0.01 by the Mann-Whitney test. ICG; indocyanine green.

Because the dilated lymphatic vessels have been related to decreased transport functionality of lymphatic vasculature, an evaluation of lymphatic vessel transport function was performed by using near-infrared (NIR) imaging system. The indocyanine green (ICG) was injected intradermally at the end of the mouse tail and NIR images were captured through side- and top-down views (**Figure 2J**). In line with the tail volume assessment, lymphatics of *S1pr1*^LECKO^ mice subjected to lymphedema surgery exhibited decreased lymphatic drainage and increased lymphatic leakage (**Figure 2K and 2L**). Together, these data suggest that loss of lymphatic S1P/S1PR1 signaling causes a more severe decline of lymphatic function and worsens lymphedematous tissue swelling. Pathways critical for lymphatic repair in disease conditions also play important roles in lymphatic development ^18^; however, assessment of LEC S1P/S1PR1 signaling during early embryonic stage illustrated only minimal dorsal edema **(Figure S6)**, indicating S1P signaling through S1PR1 is not crucial for early lymphatic development characterized by lymphangiogenesis. This study also suggests that in pathological conditions, LEC S1PR1 signaling may regulate other biological processes beyond lymphangiogenesis.

### LEC *S1pr1* deficiency increases CD4 T cell infiltration

CD4 T cells regulate lymphedema pathology by inhibiting lymphatic vessel pumping capacity, decreasing collateral lymphatic formation, and increasing lymphatic leakage ^3, 4, 36, 37^. Prior studies demonstrated that LECs regulate T cell differentiation ^11, 38, 39^. We hypothesized that LEC *S1pr1* deficiency skews pathological T cell differentiation in lymphedema. Flow cytometric analyses of single-cell suspension of lymphedematous tissues harvested from WT and *S1pr1*^LECKO^ mice were performed, and the T cell populations were assessed (**Figure 3A and S7)**. There was an increased accumulation of CD45^+^ immune cells in the tail skin of *S1pr1*^LECKO^ as compared with WT skin following lymphedema surgery (**Figure 3B**). Further analysis demonstrated that lymphedema tissue from *S1pr1*^LECKO^ mice contained significantly more CD4^+^ T cells compared with WT littermate controls, but not CD8^+^ T cells (**Figure 3C and 3D**). We focused our analysis on CD4 T cells subsets ^40–42^ and demonstrated increased IFN-ɣ-producing Th1 cells and interleukin (IL)-4-expressing Th2 cells (**Figure 3E and 3F**), and decreased Foxp3^+^CD25^+^ regulatory T cells (Tregs) (**Figure 3G**) in lymphatic *S1pr1*-deficient lymphatic mice. Recent research shows that tissue-resident memory CD4^+^ (T_RM_) cells are associated with vasculitis pathogenesis ^43, 44^ and we observed increased both CD4 T_RM_ and CD4 non-T_RM_ cell populations in *S1pr1*^LECKO^ lymphedema tail skin **(Figure S8A and S8B)**. Evaluation of IL-4-producing CD103^+^CD4^+^ T_RM_ cells demonstrated a significant increase of this population in *S1pr1*^LECKO^ lymphedema tissues **(Figure S8C through S8E)**, indicating CD4^+^ T_RM_ cells comprise an important source of CD4 T cells in a more severe form of experimental lymphedema. An IF study demonstrated increased numbers of CD4 T cells located in close proximity to lymphatic vessels in lymphedema tissue from *S1pr1*^LECKO^ mice (**Figure 3H and 3I).** These results suggest that LEC *S1pr1* deficiency may attract more CD4 T cells to lymphatic cells and promote pathological differentiation and expansion of these T cells in lymphedema.

**Figure 3.**
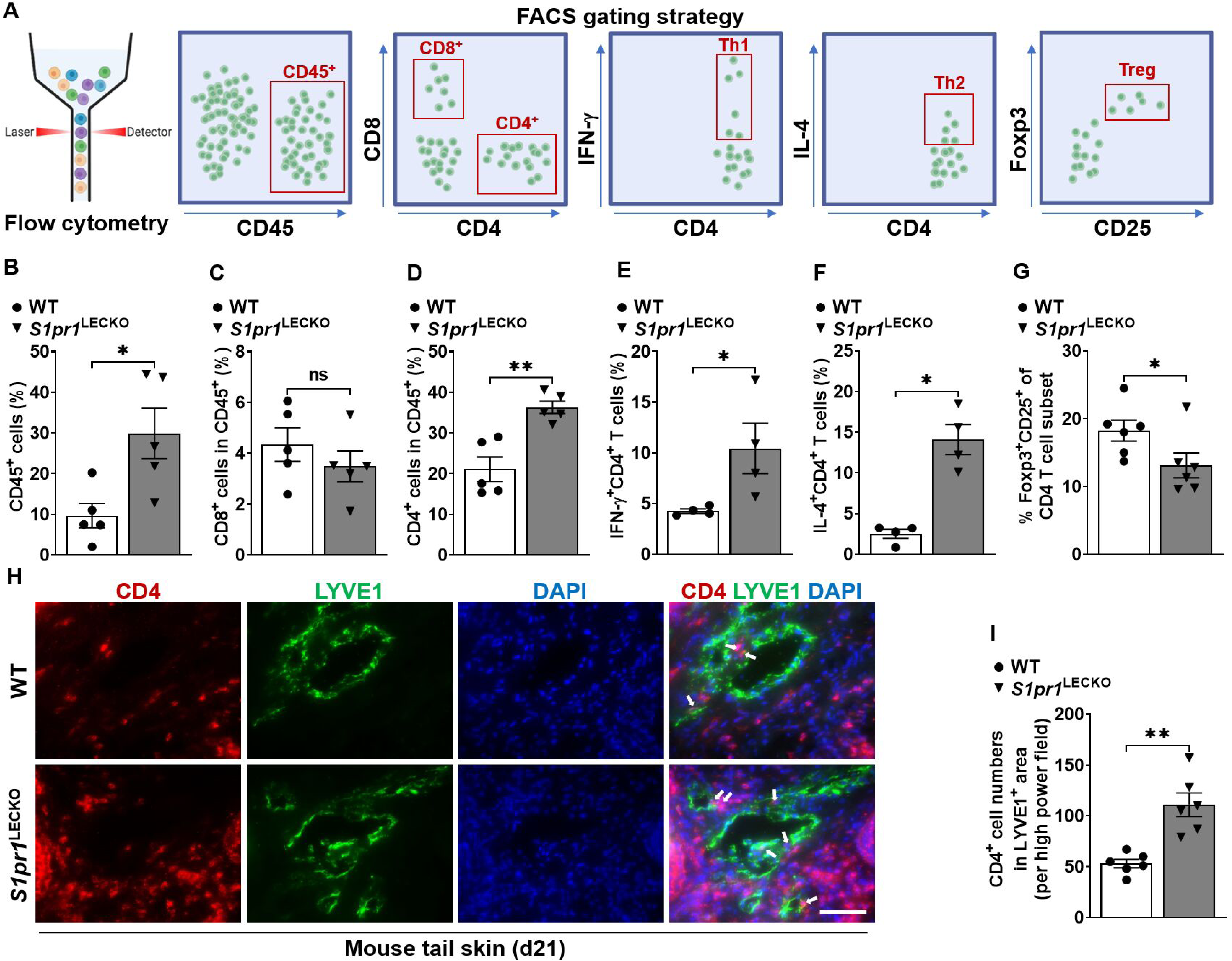
LEC *S1pr1* deficiency promotes CD4 T cell infiltration following lymphatic surgery. **(A)** Flow cytometric gating scheme for determining tail skin immune cell populations. Flow cytometric analysis was performed d21 after lymphatic surgery. **(B-G)** Quantification of CD45^+^ cells (B), CD8^+^ T cells, (C), CD4^+^ T cells (D), CD4^+^IFN-ɣ^+^ Th1 cells (E), CD4^+^IL-4^+^ Th2 cells (F), and Foxp3^+^CD25^+^CD4^+^ Treg cells (G) in tail skin (n ≥ 4 per each group). **(H)** Representative IF staining of CD4 (red) and LYVE1 (green) of the lymphedema mouse tail skin from WT or *S1pr1*^LECKO^ mice. DAPI (blue) stains for the nucleus. Arrows indicate CD4 T cells surrounding lymphatic vessels. Scale bar = 50 µm. **(I)** Quantification of the CD4 T cell staining presented in H (n ≥ 5 per each group). Data are presented as mean ± SEM; * p < 0.05, ** p < 0.01, and ns (not significant) by the Mann-Whitney test.

### LEC S1P/S1PR1 signaling regulates CD4 T cell differentiation

Emerging evidence demonstrates the LEC’s ability in regulating adaptive immune responses ^39^. To assess whether LEC S1P signaling deficiency preferentially skews CD4 T cell differentiation toward certain Th pathways, LEC-dependent CD4 T cell activation and differentiation assays were performed. Mouse lymph node-LECs (mLN-LECs) were sorted by FACS by selecting CD45(-) population that expresses both Gp38 and CD31 (**Figure 4A**). The purity of mLN-LECs was approximately 99% after sorting (**Figure 4B**). Sorted LECs were co-cultured with mouse splenocyte-derived naïve CD4 T cells in the presence of anti-CD3/28 antibodies (Ab) stimulation for 4 days (**Figure 4C**). *S1pr1*-deficient LECs significantly enhanced IFN-ɣ and IL-4 production in T cells (**Figure 4D and 4E),** while reducing the relative Foxp3^+^CD25^+^ population (**Figure 4F**). These results suggest that *S1pr1*-deficient LECs preferentially promote Th1 and Th2 effector cells and suppress Treg cell differentiation.

**Figure 4.**
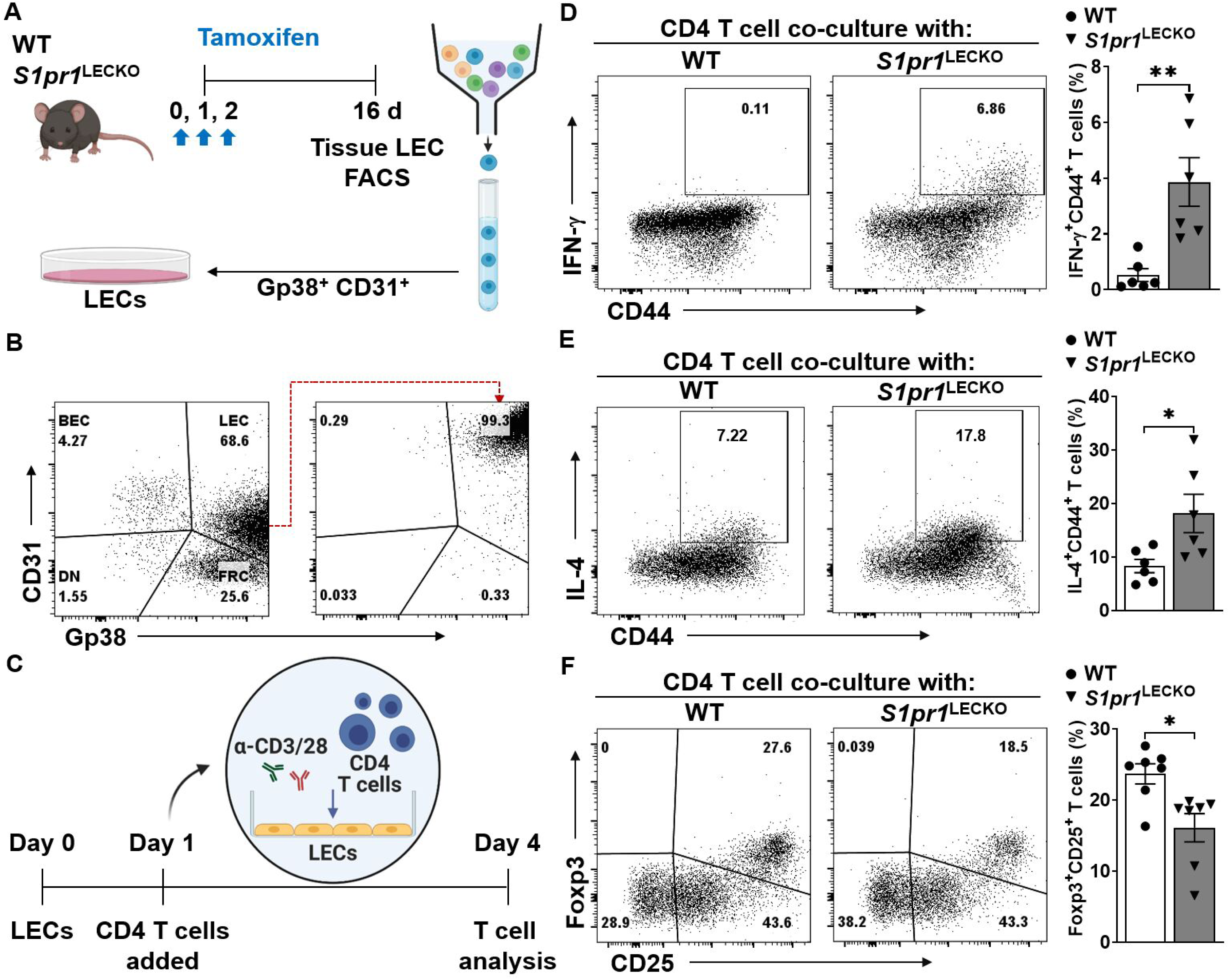
Decreased S1PR1 signaling in LECs promotes T cell activation. **(A)** Schematic diagram of LEC (Gp38^+^CD31^+^) purification. **(B)** Representative flow cytometry sorting strategy for the isolation of LECs from cultured lymph node stromal cells. **(C)** Timeline of co-culture with purified LECs and naïve CD4 T cells activating with to α-CD3/28 Abs. LECs to T cell ratio was 1:5. **(D-F)** Flow cytometric analysis was performed d4 after co-culture. Representative flow cytometric plots and quantification of IFN-ɣ^+^CD44^+^ in CD4^+^ T cells (D), IL-4^+^CD44^+^ in CD4^+^ T cells (E), and Foxp3^+^CD25^+^ in CD4^+^ T cells (F) (n ≥ 6 per each group). Data are presented as mean ± SEM; * p < 0.05 and ** p < 0.01 by the Mann-Whitney test.

Because LECs have tissue-specific tolerogenic properties ^45, 46^, we tested whether LECs from peripheral tissues also exhibit immunomodulatory function. We used lentiviral vectors that express short hairpin RNA targeting *S1PR1* (sh*S1PR1*) to knockdown S1PR1 in primary human dermal LECs (HDLECs) **(Figure S9A through S9C)**. In HDLECs, *S1PR1* expression is the highest, with S1PR4 and S1PR5 undetectable **(Figure S9D).** We then asked whether S1PR1 knockdown influences the expression of other detectable *S1PRs* and *SPHKs*. Our data demonstrated that mRNAs of *S1PR2, S1PR3, and SPHK1* were decreased in HDLECs with *S1PR1* knockdown **(Figure S9E through S9I)**. sh*Control* (sh*Ctr*)- or sh*S1PR1*-treated HDLECs (sh*S1PR1*-HDLECs) were co-cultured for three days with naïve CD4 T cells, purified from healthy individual peripheral blood mononuclear cells (PBMCs), in the presence of anti-CD3/28 Abs (**Figure 5A and 5B**). CD4 T cells, co-cultured with sh*S1PR1*-HDLECs, generally secreted higher amounts of the Th1 cytokine, IFN-ɣ, and the Th2 cytokine, IL-4, compared with T cells co-cultured with sh*Ctr-*HDLECs (**Figure 5C and 5D),** while reducing Treg (Foxp3^+^CD25^+^) population (**Figure 5E**). Multiplex cytokine measurement of CD4 T cell-related cytokines, IL-2 (T cell proliferation and activation), IFN-ɣ (Th1), and IL-4/5/13 (Th2) showed that sh*S1PR1*-treated HDLECs significantly increased the cytokine production by CD4^+^ T cells compared with sh*Ctr*-HDLECs (**Figure 5F to 5J**). Co-culture of those cells in a *trans*-well system (**Figure 5K**) indicated that the population of IFN-ɣ^+^, IL-4^+^, and Foxp3^+^ T cells was similar in sh*Ctr*- and sh*S1PR1*-HDLECs (**Figure 5L through 5N)**. Taken together, these results suggest that LECs regulate CD4 T cell differentiation via a mechanism which requires cell-cell contact.

**Figure 5.**
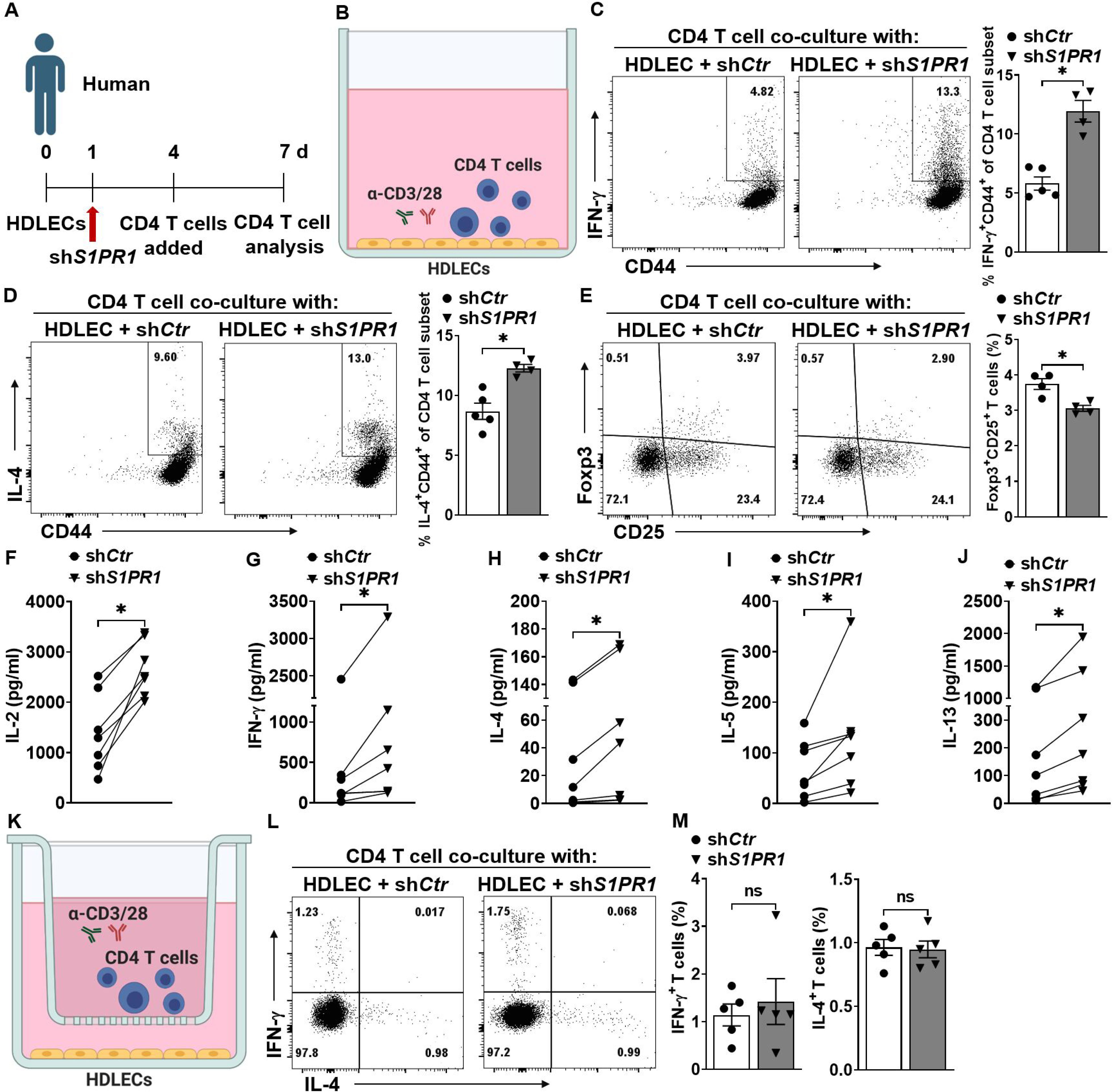
Abnormal lymphatic S1PR1 signaling activates lymphocytes through direct cell-cell contact. **(A)** Timeline of the co-culture of purified naïve CD4 T cells and HDLECs. **(B)** sh*Ctr* or sh*S1PR1*-treated HDLECs were co-cultured with purified human naïve CD4 T cells activating with α-CD3/28 Abs. LEC to T cell ratio was 1:5. **(C to E)** After 3 days of culture, human CD4 T cells were intracellularly stained for IFN-ɣ, IL-4, and Foxp3. Representative flow cytometric plots and quantification of IFN-ɣ^+^CD44^+^ in CD4^+^ T cells (C), IL-4^+^CD44^+^ in CD4^+^ T cells (D), and Foxp3^+^CD25^+^ T cells (E) (n ≥ 4 per each group). **(F to J)** T cell-related cytokines were detected in supernatant from co-cultured cells (n = 7 per each group). **(K)** HDLECs treated with sh*Ctr* or sh*SPR1* were co-cultured with human naïve CD4 T cells in 0.4 µm membrane pore trans-well in the presence of α-CD3/28 antibody for 3 days. **(L and M)** Representative flow cytometric plots (L) and compiling data (M) of intracellular IFN-ɣ^+^ and IL-4^+^ cells in the CD4^+^ T cell population cultured in a trans-well system (n ≥ 5 per each group). **(N)** Representative flow cytometric plots intracellular Foxp3^+^CD25^+^ T cells in a trans-well system. (n ≥ 4 per each group). Data are presented as mean ± SEM; * p < 0.05 and ns (not significant) by the Mann-Whitney test for C, D, and L; by Wilcoxon matched-pairs signed rank test for E, F, G, H, and I.

### Increased S1P signaling alleviates lymphedema development

Given that LEC-specific *S1pr1* deficiency significantly exacerbates lymphatic function and skews towards inflammatory CD4 immune responses, we asked whether increasing S1P signaling could restore pathological lymphatic remodeling and alleviate lymphedema. We used 4-deoxypyridoxine (4-DP), which inhibits the degradation of S1P by dampening S1P lyase activity. 10m/kg 4-DP was administered intraperitoneally (i.p.) to WT B6 mice daily following lymphedema surgery **(Figure S10A)**. 4-DP therapy reduced tail swelling **(Figure S10B and S10C)** and lymphatic remodeling **(Figure S10D and S10E)** in mice with lymphedema. Evaluation of immune cell composition in lymphedema tissue revealed significantly ameliorated Th1/2 immune responses in samples harvested from 4-DP treated mice **(Figure S10F through S10H)**. Together, our results indicate that LEC S1P signaling is vital during lymphatic repair and important for the control of local pathological immune responses in lymphedema.

### S1P signaling suppresses P-selectin expression in HDLECs

To identify putative S1P-regulated targets expressed in LECs that regulate T-cell differentiation, we performed bulk RNA-sequencing to characterize the gene expression change in HDLECs following *S1PR1* knockdown. We prepared RNA extraction from *S1PR1* knockdown HDLECs with or without CD4 T cell co-culture (**Figure 6A and 6B**). *SELP*, encoding the adhesion protein P-selectin, is the highest upregulated gene among those commonly upregulated (7 genes; *SELP, RGS5, DIRAS3, ADAMTS18, ADRA1D, APLN, IL33*) in sh*S1PR1*-HDLECs with or without CD4 T cells in the culture (**Figure 6C through 6E)**. Differential expression analysis showed a total of 18 upregulated genes in sh*S1PR1*-HDLECs without CD4 T cell co-culture and 111 upregulated in sh*S1PR1*-HDLECs with CD4 T cell co-culture **(Table S6 and S7)**. *SELP* was detected in the intersection of 7 overlapping upregulated genes between sh*S1PR1*-HDLECs with or without CD4 T cell co-culture **(Figure S11)**. Gene set enrichment analysis showed a significantly modulated pathway with up-regulated genes in sh*Ctr*-HDLECs versus sh*S1PR1*-HDLECs with CD4 T cell co-culture and suggested the upregulated genes by S1PR1 knockdown were enriched for genes involved in cell-cell contact and T cell activation (**Figure 6F and Table S8)**. Flow cytometry analysis confirmed increased cell surface P-selectin protein expression in HDLECs following *S1PR1* knockdown in sh*S1PR1* multiplicity of infection M.O.I.-dependent manner (**Figure 6G through 6I)**. Additionally, we confirmed the expression of lymphatic P-selectin dramatically increases in *S1pr1*^LECKO^ mice compared to that of the WT mice (**Figure 6 J and 6K)**.

**Figure 6.**
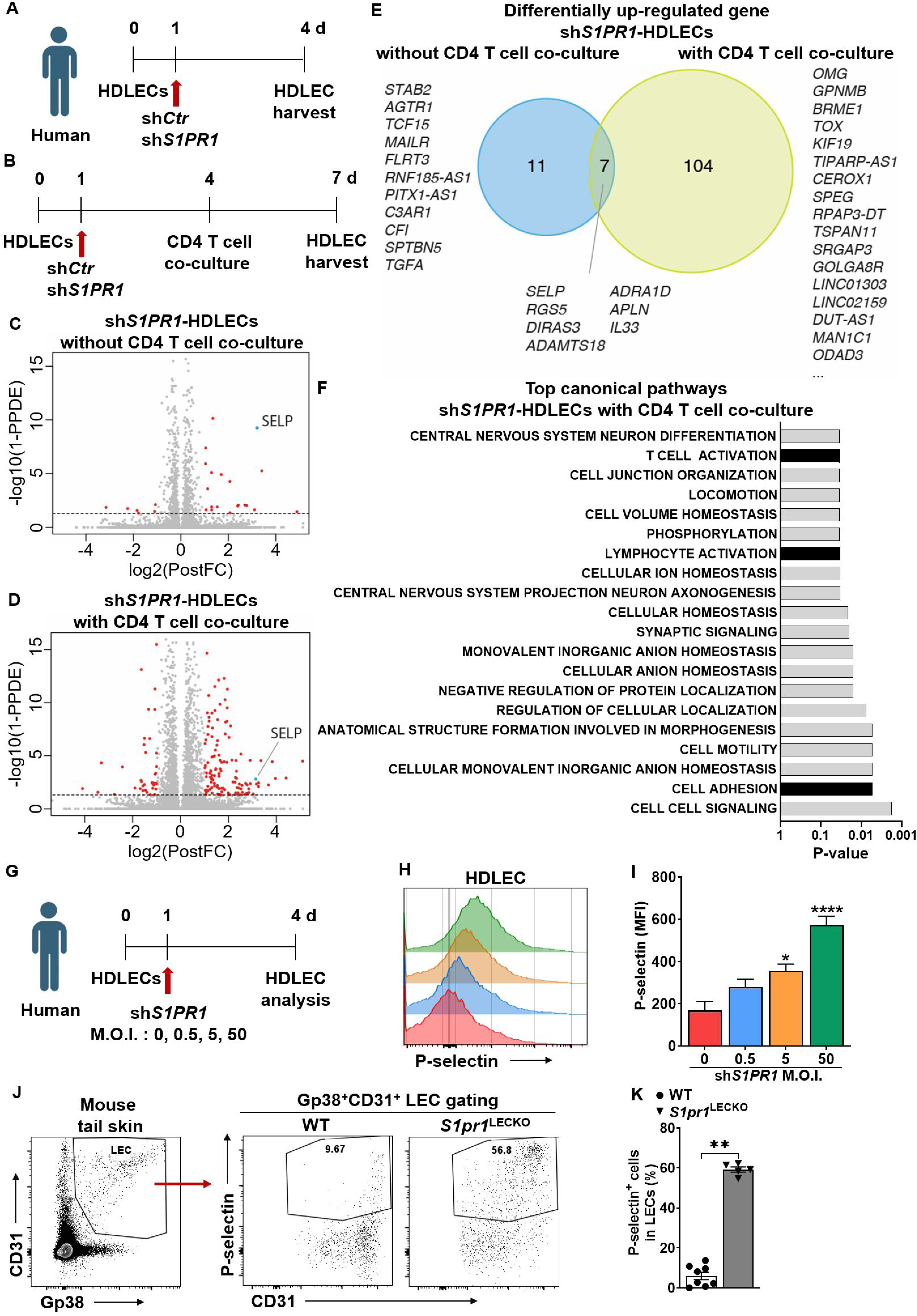
The effects of abnormal S1PR1 signaling on lymphatic biology. **(A and B)** Timeline of HDLEC treatment and harvesting time point. RNA was extracted from HDLEC without (A) or with (B) purified naïve CD4 T cell co-culture with α-CD3/28 Ab stimulation. **(C and D)** Volcano plot identifying genes significantly up-regulated (red) in sh*S1PR1*-treted HDLECs versus sh*Ctr*-treated HDLECs. (C) 18 upregulated genes from HDLECs without CD4 T cell co-culture. (D) 111 upregulated genes from HDLECs with CD4 T cell co-culture. **(E)** Venn diagram shows data summary of differentially up-regulated genes from RNA-seq data comparing sh*S1PR1*-treated HDLECs with or without CD4 T cell co-culture. Threshold of false discovery rate < 0.05. **(F)** KEGG pathway enrichment analysis of differentially up-regulated gene between sh*Ctr*-treated HDLEC and sh*S1PR1*-treated HDLEC with CD4 T cell co-culture. Most significantly upregulated pathways are shown. **(G)** Timeline of M.O.I. dependence sh*S1PR1* treatment to HDLECs. **(H and I)** P-selectin fluorescence intensity in HDLECs after *S1PR1* knock-down was evaluated by flow cytometric analysis. Representative (H) and compiling data (I) are shown (n ≥ 4 per each group). **(J and K)** Flow cytometric analysis was performed from tail skin of WT and *S1pr1*^LECKO^ mice. Representative flow cytometric plots (J) and quantification of P-selectin^+^ LECs in tail tissue skin (K) (n ≥ 4 per each group). Data for I are presented as the mean ± SEM; * p < 0.05 and **** p < 0.0001 compared with the sh*Ctr*-treated HDLECs group; by Ordinary one-way ANOVA. Data for K are presented as the mean ± SEM; ** p < 0.01 compared with the WT group; by Mann-Whitney test. KEGG; Kyoto Encyclopedia of Genes and Genomes. M.O.I.; multiplicity of infection.

### P-selectin blockade inhibits the activation of CD4 T cells co-cultured with *S1PR1*-deficient HDLECs

Cutaneous lymphocyte-associated antigen (CLA) is a carbohydrate modification form of P-selectin glycoprotein ligand-1 (PSGL-1), which is a high affinity receptor for P-selectin and its expression is up-regulated during T cell activation ^47, 48^. These studies suggested that P-selectin engagement of PSGL-1/CLA expressed on T cells may regulate T cell differentiation. Consistent with published studies, the CLA^+^ CD4 T cell population increased following 3 days of culture in the presence of anti-CD3/28 Abs **(Figure S12A)**. Blocking PSGL-1/CLA signaling with P-selectin-specific antibody, Waps 12.2, suppressed IFN-γ and IL-4 expression in CD4 T cells co-cultured with sh*S1PR1-*treated HDLECs in an antibody dose-dependent manner (**Figure 7A through 7C)**. These data suggest that S1P signaling deficiency induces P-selectin expression which, in turn, mediates enhanced Th1 and Th2 CD4 T cell differentiation.

**Figure 7.**
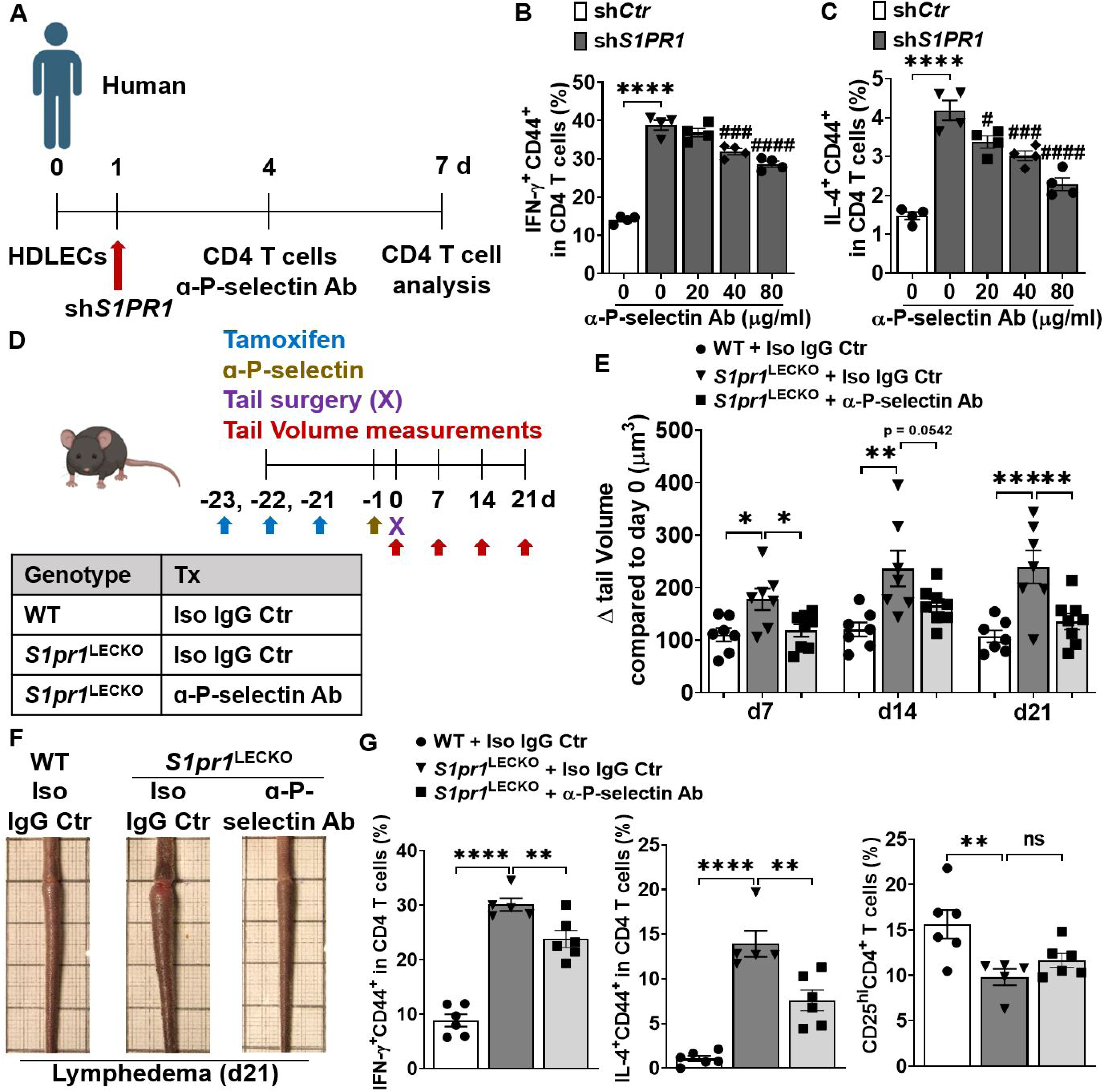
Blocking P-selectin decreases CD4 T cell activation and lymphedema. (**A**) Timeline of the co-culture of purified naïve CD4 T cells and HDLEC. ɑ-human P-selectin Ab (Waps12.2) was added to sh*S1PR1*-treated HDLECs 1h before purified memory CD4 T cell co-culture with HDLECs at day 4. **(B and C)** Flow cytometric analysis was performed d3 after co-culture. Quantification of IFN-ɣ^+^CD44^+^ in CD4^+^ T cells (B), IL-4^+^CD44^+^ in CD4^+^ T cells (C) (n = 4 per each group). **(D)** Schematic diagram of the experimental protocol. 5 mg/kg anti-mouse P-selectin Ab (RB40.34.4) or Isotype IgG control (Iso IgG Ctr) was retro-orbital-i.v. injected into *S1pr1*^LECKO^ mice 1 day before lymphedema surgery and the tail size of animals was measured at days 0, 7, 14, and 21. **(E and F)** Quantification of tail volume changes (E) (n ≥ 7 per each group). Representative photographs of tail skin on day 21 after surgery (F). **(G)** Quantification of IFN-ɣ^+^CD44^+^, IL-4^+^CD44^+^, and CD25^hi^ in CD4^+^ T cells from tail skin 21 day after lymphedema surgery (n ≥ 5 per each group). Data B and C are presented as the mean ± SEM; **** p < 0.0001, # p < 0.05, ### p < 0.001, and #### p < 0.0001 compared with the sh*S1PR1*-treated HDLEC group; by Ordinary one-way ANOVA. Data in E and G are presented as mean ± SEM; * p < 0.05, ** p < 0.01, *** p < 0.001, **** p < 0.0001, and ns (not significant) compared with the *S1pr1*^LECKO^ + Iso Ctr group; by Ordinary one-way ANOVA.

### Anti-P-selectin Ab alleviates lymphedema developed in *S1pr1*^LECKO^ mice

To assess the relevance of P-selectin in lymphedema, we evaluated the effect of blocking P-selectin in lymphedema pathogenesis. 5 mg/kg anti-mouse P-selectin Ab (RB40.34.4) ^49^ was injected intravenously (i.v.; retro-orbital) into *S1pr1*^LECKO^ mice one day before lymphedema surgery, and tail volumes were monitored weekly (**Figure 7D**). *S1pr1*^LECKO^ mice treated with RB40.34.4 showed a significant decrease in tail swelling compared to *S1pr1*^LECKO^ mice treated with control IgG (**Figure 7E and 7F**). A flow cytometry analysis of CD4 T cell populations in lymphedematous tail skin demonstrated significant decrease of IFN-ɣ and IL-4-producing CD4 T cells in samples derived from mice treated with anti-P-selectin (**Figure 7G**). Although not significant, Treg (CD25^hi^ CD4^+^) population showed a trend of increase in lymphedema mice treated with RB40.34.4 comparing to isotype-treated subjects (Figure 7G). Additionally, lymphedema skin from anti-P-selectin Ab-treated mice contained significantly reduced CLA-expressing CD44^+^ CD4 T cells **(Figure S12B and S12C)**. To further evaluate the therapeutic potential of anti-P-selectin, RB40.34.4 was administered 2 days following lymphedema surgery, when tails already developed noticeable swelling. Our data illustrated a trend of tail swelling reduction in lymphedema mice treated with anti-P-selectin antibody comparing to isotype-treated mice **(Figure S13)**. Collectively, these data suggest that P-selectin blockade improves lymphatic tissue swelling and pathogenic CD4 T cell accumulation in severe lymphedema.

## Discussion

The lymphatic vasculature is an integral circulatory system and crucially influences disease pathophysiology ^7, 50–59^. Lymphedema is the prototypical lymphatic vascular disorder that afflicts nearly 200 million individuals globally with no approved drugs ^60^. A better understanding of lymphedema pathobiology can identify much-needed therapeutic targets. While inflammation is increasingly recognized as a disease-promoting pathology in lymphedema ^61^, how uncontrolled abnormal immune responses develop in this disease remains poorly understood. In this study, we showed that impaired S1P signaling in LECs promotes pathogenic CD4 T cell differentiation and exacerbates lymphatic malfunction in lymphedema. These results illustrate how pathological LEC and T cell interactions can contribute to lymphedema progression and how suppressing these intercellular relationships may attenuate the disease.

We first assessed key molecules involved in the S1P signaling in lymphedema conditions. LC/MS analysis demonstrated significantly decreased S1P concentrations in both clinical and preclinical serum samples. Our clinical data also support that serum S1P concentrations are inversely associated with age; notably, menopause as a factor reducing S1P levels ^62^. Body mass index (BMI) and metabolic status may also influence S1P concentrations, especially in diabetic patients^63^. However, how BMI may affect S1P levels in our cohort is unclear because relevant information for control subjects is lacking.

Evaluation of SPHK1, the main SPHK isoform, indicated a diminished expression of this protein in LECs, which is the major S1P producer in peripheral tissues ^34, 35^. These data suggest that lymphedema tissues have reduced local S1P production concordant with reduced systemic levels. Additionally, lymphatic vessels in lymphedema tissue exhibited low expression levels of the main isoform of S1P receptors, S1PR1. While our data clearly demonstrate that decreased S1P production may result from diminished SPHK1 expression in LECs, we do not exclude the possibility that reduced SPHK1 in other cell types may also contribute to S1P reduction in lymphedema. Future single cell RNA-Seq may provide more comprehensive information on SPHK1 expression in different cell types in lymphedema and offer additional insights.

We previously demonstrated that the proinflammatory lipid mediator, LTB_4_, produced chiefly by macrophages, suppresses SPHK1 expression in blood endothelial cells ^64^ . Because LTB_4_ is pathogenic in lymphedema ^65^, LTB_4_ may similarly reduce SPHK1 expression in LECs. A recent study shows that inflammation-mediated autophagy induces the degradation of SPHK1, resulting in diminished S1P production in LECs ^66^, suggesting the inflammation is generally sufficient to reduce SPHK1 levels. Additionally, LTB_4_ suppresses LEC Notch activity ^65^; and Notch signaling sustains S1PR1 expression ^67^. Thus, increased LTB_4_ production in lymphedema may also reduce LEC S1PR1 expression. LTB_4_ antagonism for upper extremity lymphedema is currently being evaluated in a Phase II clinical trial (NCT05203835). The data from the current study suggest that lymphedema has diminished lymphatic vessel S1P signaling characterized by the decrease of both the ligand (S1P) production and receptor (S1PR1) expression, possibly as a result of increased levels of proinflammatory mediators generated by cells of the innate immunity, such as macrophages.

To evaluate the biological effects of diminished LEC S1P signaling in lymphedema, we generated the LEC-specific *S1pr1* loss-of-function mice (*S1pr1*^LECKO^) and found that *S1pr1*^LECKO^ mice develop more severe lymphedema with exacerbated tail swelling, cutaneous thickening, increased lymphatic leakage, and aggravated tissue inflammation. Conversely, enhanced S1P signaling by systemic 4-DP treatment improved tail swelling, lymphatic vascular function, and inflammation. Together, these data indicate that LEC S1P signaling through S1PR1 is crucial for lymphatic vascular repair and the control of pathological tissue inflammation.

While existing data from both clinical and preclinical studies reveal exuberant inflammation characterized by dysregulated adaptive immune cells infiltration in lymphedema tissues ^3, 37^, how malfunctioning adaptive immune responses propagate is not completely understood ^39^. We first compared immune cell compositions in the skin tissue of lymphedema of WT vs *S1pr1*^LECKO^ mice. Our data showed significantly increased Th1 (IFN-γ^+^CD4^+^) and Th2 (IL-4^+^CD4^+^) cells but decreased immunosuppressive Treg cells (CD4^+^CD25^+^FoxP3^+^) in *S1pr1*^LECKO^ mice comparing to that observed in controls. Because Th1 and Th2 cells are pathogenic in lymphedema, whereas Treg cells are demonstrably protective ^36, 68^, our data suggest that LEC *S1pr1* deficiency exacerbates pathogenic inflammatory responses in lymphedema.

Additionally, our immune profiling result demonstrated a significantly increased CD69^+^CD103^+^ [markers for Tissue-resident memory cells (T_RM_) ^69–71^], IL-4-producing CD4 T cell population in lymphedema tissue from *S1pr1*^LECKO^ mice, suggesting that LEC *S1pr1* deficiency promotes the expansion of T_RM_ population in lymphedema. T_RM_ cells play critical roles in tissue-specific immune and inflammatory diseases ^72, 73^ and are associated with vasculitis ^43, 44^. LEC *S1pr1* deficiency-associated CD4^+^ T_RM_ expansion may play an important pathogenic role in lymphedema progression. Interestingly, several studies have demonstrated that Th2 immune responses are more critical than Th1 immune responses in lymphedema pathogenesis ^4, 5, 74^, which appear to support our observation that the dominancy of IL-4^+^CD4^+^ T_RM_ cells over IFN-ɣ^+^CD4^+^ T_RM_ cells in lymphedema tail skin of *S1pr1*^LECKO^ mice.

Because LECs can directly regulate T cell immunity and are important immunomodulators of peripheral tolerance ^10, 39, 75, 76^, we went on to ask whether LECs with deficient S1P signaling skew T cell differentiation and enhance inflammation in lymphedema. *S1pr1*-deficient LECs isolated from LEC-specific *S1pr1* KO mice promoted mouse Th1/Th2 cell differentiation but decreased the immunosuppressive Treg cell subset. Consistently, co-culture of naïve human CD4^+^ T cells with sh*Ctr*- or sh*S1PR1*-HDLECs illustrated that human LECs with *S1PR1* knockdown similarly enhance human Th1/Th2 T cell differentiation. Using the trans-well system, we further show that LEC-modulated skewed T cell differentiation requires cell-cell contact. Collectively, our data suggests that the loss of S1P signaling may lead to altered expression of certain cell surface proteins, which in turn, affects local T cell differentiation. S1P directly acts on T cells to promote Treg differentiation ^77^, a mechanism that may partly explain the reduction Treg population in *S1pr1*^LECKO^ mice. While previous experiments suggested that lymphatic damage-induced CD4 T cell immunity involves dendritic cell (DC) migration to the draining lymph node, immune priming, and T cell trafficking back to the local lymphedematous tissue ^36^, our data suggests that in lymphedema, activation of the pathogenic adaptive immunity may be enhanced by S1P signaling-deficient local LECs.

To find genes involved in T cell reprogramming by S1P signaling-deficient LECs, we performed bulk RNA-seq analyses. Among the most significantly changed pathways is the cell adhesion signaling and upregulation of the adhesion molecule P-selectin ^78^. Our data appear to be consistent with findings from blood vessel endothelial cells in which inflammation-induced S1PR1 deficiency promotes P-selectin expression and enhances leukocyte extravasation ^79, 80^. Blocking lymphatic P-selectin signaling reversed the effect of S1P signaling deficiency in promoting Th1/Th2 differentiation in vitro; P-selectin neutralization significantly alleviated tail swelling and tissue inflammation in severe lymphedema. These results demonstrate that the loss of LEC S1P signaling may induce a proinflammatory LEC phenotype with increased expression of adhesion molecules such as P-selectin which, in turn, influences T cell activation and differentiation. While P-selectin is best known for its role in T cell rolling during immune cell extravasation ^81, 82^, recent data indicated that P-selectin may modulate T cell activity through PSGL-1^83^. In agreement, our data suggested that increased P-selectin expression in *S1pr1*-deficient LECs can promote CD4 T cell accumulation around lymphatic vessels and directly modulate their differentiation and activation; a process that could promote abnormal immune responses in lymphedema. Targeting P-selectin is a promising strategy in treating several inflammatory diseases, such as sickle cell disease and neuroblastoma tumors ^84, 85^. Blocking P-selectin may also represent a promising new approach for treating lymphedema involving T cell immunity. Additionally, if this biology patterns human responses, it is possible that P-Selectin-directed therapy may benefit patients at risk for lymphedema progression, such as in the peri-operative period following breast cancer surgery.

Our study has several limitations. Treatments testing 4-DP and ɑ-P-selectin Ab in mice may have also targeted non-lymphatic tissues. To more definitively delineate roles of diminished LEC S1P signaling and increased LEC P-selectin expression in lymphedema, further studies require LEC-specific S1PR1 overexpression or P-selectin knockout models. Future investigations can determine local S1P levels in lymphedema tissue to better evaluate the status of ligand production in this disease. A prior study demonstrated that LEC S1PR1 deficiency delays macrophage clearance following myocardial infarction ^86^, and future assessments may prove a similar mechanism also operates in lymphedema pathogenesis.

In summary, our findings provide evidence that abnormal lymphatic S1PR1 signaling, caused by lymphatic injury, promotes pathological CD4 T cell activation, and exacerbates lymphedema progression (**Figure 8**). The evolution of lymphedema is complicated with many cellular interactions and disease mediators changing over an expanded period and in a dynamic manner. For this condition, there are a number of pathological features (e.g., inflammation, fibrosis, adiposity, vascular disease) and clinical endpoints (e.g., limb volume, skin thickness) which can be the focus of new drug and biophysical interventions. In the current effort, we have suggested that reduced S1P signaling in lymphatics may presage the onset of lymphedema and is therefore a potentially useful biomarker of disease risk. The use of P-selectin inhibitors for lymphedema is a promising consideration for future clinical trials.

**Figure 8.**
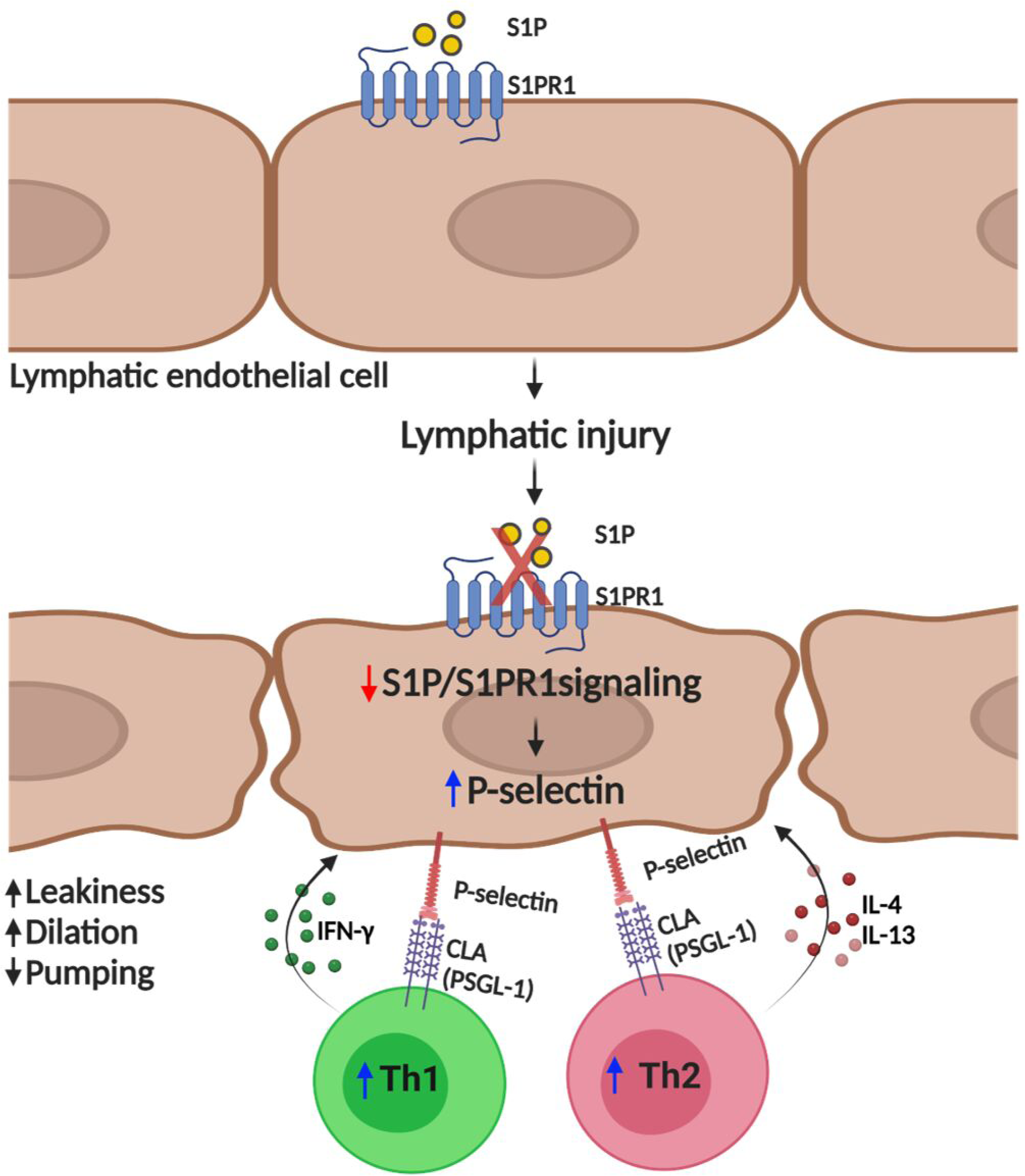
LEC S1PR1 signaling in lymphedema pathogenesis. Graphic abstract showing how abnormal lymphatic endothelial cell (LEC) S1P signaling can induce T cell activation and lymphedema after lymphatic injury. Lymphatic injury caused LEC S1P-S1PR1 signaling reduction. These changes collectively induce P-selectin expression on LECs resulting in CD4 T cell overactivation.

## Data Availability

We will adhere to the current NHLBI and NIH Data Management and Sharing Policy (NOT-OD-21-013) on sharing of the results and accomplishments of activities, making these available to the research community and to the public at large.

## Acknowledgments

Author contribution: DK, WT, and XJ planned and performed experiments and were responsible for data analysis. RV, JC, SG, SL, TG, SK, YZ, ECS, and EB performed the experiments. TW and MX analyzed the RNA-seq data. JBD, PK, JP, and SR provided intellectual input and reagents. DK, WT, XJ, and MRN conceived the study and wrote the manuscript.

## Abbreviations

LEC: Lymphatic endothelial cell
S1P: Sphingosine-1-phosphate
SPHK1: Sphingosine kinase 1
S1PR1: S1P receptor 1
APC: Antigen-presenting cell
LN: Lymph node
TRM: Tissue-resident memory T
Th1/2: T helper type 1/2 cells
Tregs: Regulator T cells
LYVE1: Lymphatic vessel endothelial receptor 1
Prox1: Prospero homeobox 1 (Prox1)
NIR: Near-infrared
HDLEC: Human dermal LEC
FACS: Fluorescence-activated cell sorting
WT: Wild type
S1pr1LECKO: Lymphatic endothelial cell-specific S1pr1 knockout
IL: Interleukin
IFN: Interferon
4-DP: 4-deoxypyridoxine
CLA: Cutaneous lymphocyte-associated antigen
PSGL-1: P-selectin glycoprotein ligand-1

